# The impact of COVID-19 on patients with asthma

**DOI:** 10.1101/2020.07.24.20161596

**Authors:** José Luis Izquierdo, Carlos Almonacid, Yolanda González, Carlos Del Rio-Bermúdez, Julio Ancochea, Remedios Cárdenas, Joan B Soriano

## Abstract

**Background:** From the onset of the COVID-19 pandemic, an association between the severity of COVID-19 and the presence of certain medical chronic conditions has been suggested. However, unlike influenza and other viruses, the burden of the disease in patients with asthma has been less evident.

**Objective:** This study aims at a better understanding of the burden of COVID-19 in patients with asthma and the impact of asthma, its related comorbidities, and treatment on the prognosis of COVID-19.

**Methods:** We analyzed clinical data from patients with asthma from January 1^st^ to May 10^th^, 2020 using big data analytics and artificial intelligence through the SAVANA *Manager*^®^ clinical platform.

**Results:** Out of 71,192 patients with asthma, 1,006 (1.41%) suffered from COVID-19. Compared to asthmatic individuals without COVID-19, patients with asthma and COVID-19 were significantly older (55 vs. 42 years), predominantly female (66% vs. 59%), had higher prevalence of hypertension, dyslipidemias, diabetes, and obesity, and smoked more frequently. Contrarily, allergy-related factors such as rhinitis and eczema were less frequent in asthmatic patients with COVID-19 (*P* < .001). Higher prevalence of hypertension, dyslipidemia, diabetes, and obesity was also confirmed in those patients with asthma and COVID-19 who required hospital admission. The percentage of individuals using inhaled corticosteroids (ICS) was lower in patients who required hospitalization due to COVID-19, as compared to non-hospitalized patients (48.3% vs. 61.5%; OR: 0.58: 95% CI 0.44 - 0.77). During the study period, 865 (1.21%) patients with asthma were being treated with biologics. Although these patients showed increased severity and more comorbidities at the ear, nose, and throat (ENT) level, their hospital admission rates due to COVID-19 were relatively low (0.23%). COVID-19 increased inpatient mortality in asthmatic patients (2.29% vs 0.54%; OR 2.29: 95% CI 4.35 – 6.66).

**Conclusion:** Our results indicate that the number of COVID-19 cases in patients with asthma has been low, although higher than the observed in the general population. Patients with asthma and COVID-19 were older and were at increased risk due to comorbidity-related factors. ICS and biologics are generally safe and may be associated with a protective effect against severe COVID-19 infection.

## INTRODUCTION

Asthma remains a global major challenge for public health. Affecting approximately 272 million people of all ages (4.5% of adults aged 18-50), asthma is one of the most common chronic disorders worldwide (1,2). In the United States, it has been estimated that up to 12 million people experience an acute exacerbation of their asthma ever year, a quarter of which require hospitalization (3). In Europe, asthma ranks 14^th^ in terms of duration and associated disability, and leads to an estimated cost of EUR 25 billion per year (4). In Spain, 5% of adults and 10% of children have asthma. However, up to 50% of patients remain undiagnosed. Indeed, 8.6% of adults (aged 18-70) and 14% of children show asthma-related symptoms, being dyspnea and cough the most common (4,5). Yet, between 60% and 70% of patients with asthma in Spain are not properly controlled (6, 7). This is critical since patients with uncontrolled asthma may generate ten times as much direct and indirect costs than controlled patients (1).

The severity of the disease is related to uncontrolled bronchial asthma in many patients; in other cases, worsening symptoms and asthma exacerbation are associated with treatment-related variables (e.g., inappropriate treatment, lack of adherence) or with the presence of underlying risk factors (6-8), most notably viral respiratory infection. Asthma exacerbations caused by respiratory infections or other conditions have a negative impact on the patient’s health status and lead to worse prognosis.

COVID-19 is the disease caused by the severe acute respiratory syndrome coronavirus 2 (SARS-CoV-2). Clinically, the severity of COVID-19 can vary from mild to very severe, causing mortality in some patients (9,10). The ongoing COVID-19 pandemic certainly represents a major challenge for health systems globally. Since the beginning of the pandemic, an association between COVID-19 severity and chronic medical conditions such as cardiovascular disease, diabetes mellitus, and high blood pressure has been suggested. Unlike influenza and other seasonal viruses however, the impact of COVID-19 in patients with asthma has been less evident (11-14). On the other hand, the severity and mortality of COVID-19 has been strongly related with age. Although the virus can infect individuals of all ages, most severe cases to date have been described in adults 55 years and older, and in patients with the aforementioned comorbidities. In this age group, patients typically have more than one chronic condition, particularly endocrine-metabolic and cardiovascular diseases. With this background, it is imperative to characterize the clinical course of SARS-CoV-2 infection in patients with asthma and assess the impact of asthma and asthma-related comorbidities and treatment in COVID-19-related outcomes.

Emerging and rapidly evolving diseases such as COVID-19 are best understood using population-based registries containing real-world information (15). In this context, combining real-world data with big data analytics has the potential to increase our understanding of the effects of COVID-19 in patients with asthma and identify new strategies and management options for therapeutic intervention. A relevant data source with the above-mentioned characteristics is the clinical information contained in patients’ electronic health records (EHR).

Using the clinical information captured in the EHRs of patients with asthma and COVID-19, here we aimed to *a)* describe the frequency and clinical characteristics of these patients and *b)* understand the clinical impact of COVID-19 on the clinical course of patients with asthma. To achieve the study objectives, we used big data analytics and artificial intelligence (AI) through the SAVANA *Manager*^®^ clinical platform (16-17).

## METHODS

This was a multi-center, non-interventional, retrospective study using free-text data captured in the EHRs of patients diagnosed of COVID-19. The study period was January 1, 2019 - May 10, 2020.

We followed the Strengthening the Reporting of Observational Studies in Epidemiology (STROBE) guidelines for reporting observational studies (18). The study was conducted in accordance with legal and regulatory requirements and followed research practices described in the ICH Guidelines for Good Clinical Practice, the Declaration of Helsinki in its latest edition, the Guidelines for Good Pharmacoepidemiology Practice (GPP), and local regulations. Given the retrospective and observational nature of the study, physicians’ prescribing habits and patient assignment to a specific therapeutic strategy were solely determined by the physician, team, or hospital concerned. Likewise, a standard informed consent does not apply to this study. All actions towards data protection were taken in accordance with the European Data Protection authorities’ code of good practice regarding big data projects and the European General Data Protection Regulation (GDPR). This study was approved by the Ethics and Research Committee of the University Hospital of Guadalajara (Spain).

Clinical data from a total of 2,034,921 patients with available EHRs in the region of Castilla La-Mancha (Spain) were explored. Data were collected from all available services, including inpatient and outpatient departments, emergency room, and primary care.

The information from EHRs was extracted with Natural Language Processing (NLP) and AI techniques using SAVANA *Manager*^®^, a powerful multilingual (natural language) engine for the analysis of free-text clinical information. This software is able to interpret any content included in electronic clinical records, regardless of the electronic system in which it operates. Importantly, this tool can capture numerical values and clinical notes and transform them into accessible variables, thus allowing for the reuse of the information captured in large-scale collections of clinical records (i.e., big data). The data extraction process has four distinct phases aimed at transferring and aggregating the data into the study database, namely *a)* Acquisition: data acquisition is the responsibility of the participating site, in close collaboration with SAVANA’s Information Technology staff. In compliance with the EU GDPR, data were anonymized and transferred to SAVANA during this phase; *b)* Integration: In this phase, data were integrated into the database; *c)* NLP processing: SAVANA’s *EHRead*^*®*^ technology applied NLP techniques to analyze and extract the unstructured free-text information written in large amounts of EHRs. The NLP output is a synthetic patient database, as the software creates a patient database from scratch. This process ensures that this information is inaccessible and makes traceability to individual patients impossible; and *d)* Validation, consisting of a medical validation of the tool’s output by doctors and researchers.

The terminology used by SAVANA is based on multiple sources such as SNOMED CT (19), which includes medical codes, concepts, synonyms, and definitions regarding symptoms, diagnoses, body structures, and substances commonly used in clinical documentation. Due to the novel methodological approach of this study, we complemented our clinical findings with the assessment of *EHRead*’s performance. This evaluation was aimed at verifying the system’s accuracy in identifying records that contain mentions of asthma and COVID-19 and its related variables. For a comprehensive description of the evaluation procedure, see (19). Briefly, the annotations made by the medical experts were used to generate the gold standard to assess the performance of *EHRead*’s output; performance is calculated in terms of the standard metrics of accuracy (P), recall (R), and their harmonic mean F-score (20).

All statistical analyses were conducted using SPSS software (V25.0). Unless otherwise indicated, qualitative variables are expressed as absolute frequencies and percentages, while quantitative variables are expressed as means and standard deviations. For the assessment of statistical significance of numerical variables, we used independent samples Student’s T-tests or ANOVA. To measure the relative distribution of patients assigned to different categories of qualitative variables, we used Chi^2^ tests. In all cases, a p value for statistical significance was set at 0.05.

## RESULTS

We identified 71,192 patients with asthma during the study period (January 1, 2019 - May 10, 2020). The search terms used to identify patients with bronchial asthma are listed in **eTable 1**. For the linguistic evaluation of the variable ‘asthma’, we obtained Precision, Recall, and F-Scores of 0.88, 0.75, and 0.81, respectively; these metrics indicate that patients with asthma were properly identified within the target population. The patient flowchart for asthmatics with and without COVID-19 is depicted in **Figure 1**.

**Table 1.** Demographic and clinical characteristics of patients with asthma with/without COVID-19

|  | Asthma<br>(n=71,192) | Asthma without<br>COVID-19<br>(n=70,544) | Asthma with<br>COVID-19<br>(n=1,006) | OR (95% CI)<br>*P* value * |
| --- | --- | --- | --- | --- |
| *Age* |  |  |  |  |
| Years<br>(mean ± SD) | 42 ± 24 | 41 ± 24 | 55 ± 20 | < .001 |
| *Sex* |  |  |  |  |
| Female (%) | 59 | 58 | 66 | 1.35 (1.18 – 1.54)<br><.001 |
| *Tobacco use (%)* | 10 | 9 | 11 | 1.25 (1.03 -1.53)<br>0.02 |
| *Comorbidities* |  |  |  |  |
| Arterial<br>Hypertension<br>(%) | 24 | 23 | 39 | 2.02 (1.78 - 2.30)<br><.001 |
| Dyslipidemia<br>(%) | 17 | 16 | 26 | 1.85 (1.60 – 2.13)<br><.001 |
| Diabetes<br>Mellitus (%) | 11 | 11 | 16 | 1.54 (1.30 – 1.82)<br><.001 |
| Obesity (%) | 10 | 9 | 13 | 1.35 (1.12-1.62)<br><.001 |
| Rhinitis (%) | 33 | 33 | 17 | 0.42 (0.35 -0.49)<br><.001 |
| Eczema (%) | 7 | 7 | 1 | 0.50 (0.40 – 0.76)<br><.001 |

\* Patients with asthma with COVID-19 vs. patients with asthma without COVID-19

**FIGURE 1.**
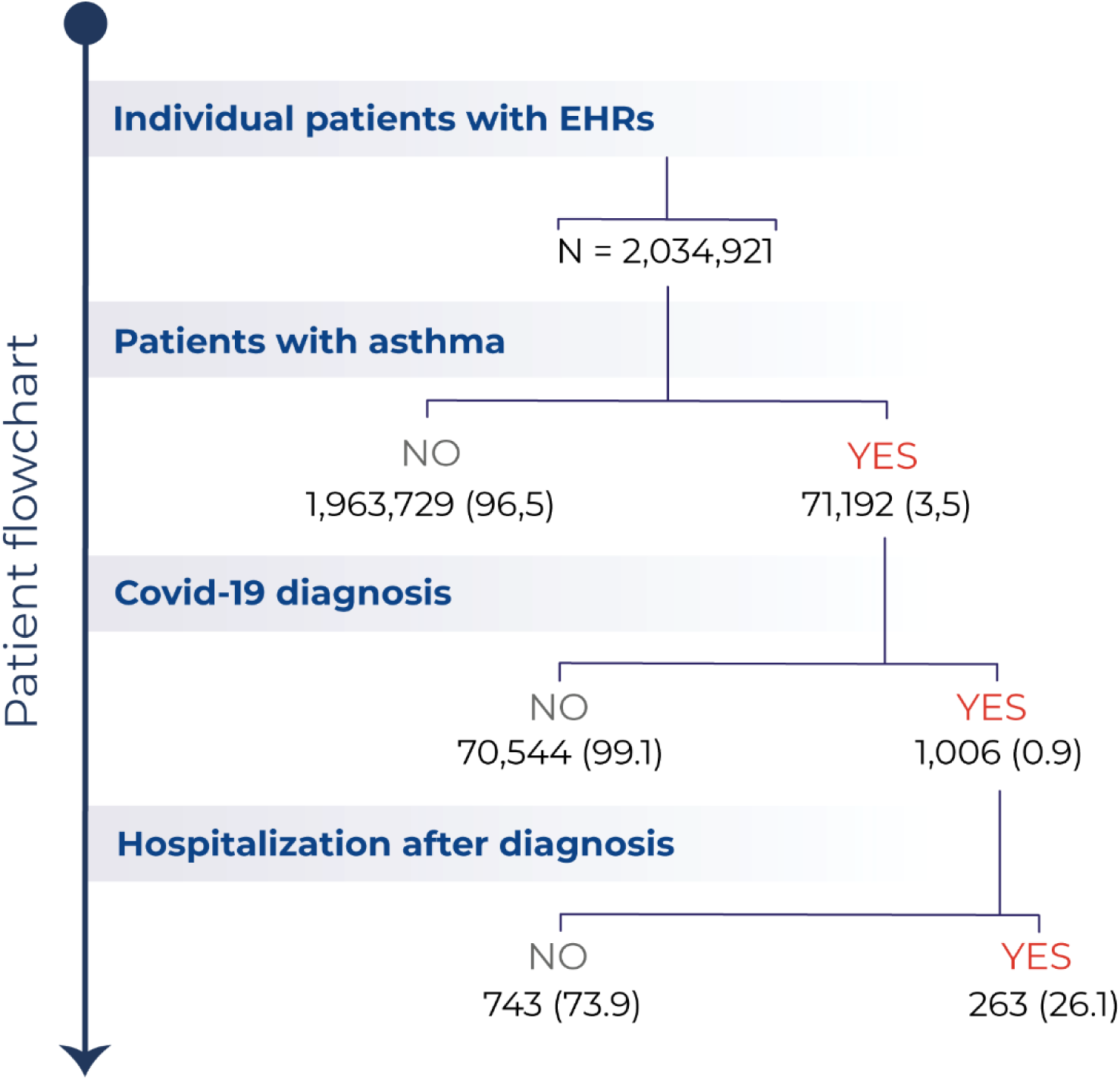
Patient Flowchart. Flowchart depicting the total number (%) of patients with available EHRs, the number of patients with asthma, the number of patients diagnosed with COVID-19, and of those, the number of hospitalizations after diagnosis during the study period (January 1^st^, 2020-May 10^th^, 2020). All percent values are computed in relation to the immediately above level.

Patients’ age (mean ± SD) was 42 ± 20 years; 59% of patients were women. Overall, 1,006 (1.41%) asthma patients were also diagnosed with COVID-19. *EHRead* identified COVID-19 with a Precision of 0,99, a Recall of 0,75, and a F-Score of 0,93; again, these results indicate that within our population with asthma, COVID-19 cases were accurately identified. COVID-19 diagnosis was confirmed by PCR in 61% of patients (n = 611); in the remaining cases, and considering the epidemiological context of the pandemic in the study area between March and May 2020, diagnosis was based on rapid serological tests or clinical, radiological, and/or analytical evaluation, Notably, the percentage (95% CI) of patients diagnosed with COVID-19 in the population of patients with asthma (1.41%; 1.33 – 1.50) was significantly higher than in the general population of Castilla La-Mancha (Spain) (0.86%; 0.85 – 0.87), *P* <.001.

Patients with asthma who also had a diagnosis of COVID-19 were older, predominantly women, and had higher prevalence rates of hypertension, dyslipidemia, diabetes, obesity, and smoking habits than asthmatic individuals without COVID-19 (all *P* <.05). By contrast, atopy-related factors such as rhinitis or eczema were significantly more frequent in patients without COVID-19 (**Table 1**). The higher prevalence of hypertension, dyslipidemia, diabetes, and obesity was further confirmed in those patients requiring hospital admission, as compared with those who only required outpatient management (**Table 2**).

**Table 2.** Demographic and clinical characteristics of patients with asthma and COVID-19 that required a hospital admission.

|  | Asthma +<br>COVID-19<br>(n=1,006) | Asthma + COVID-19<br>Non-Hospitalized<br>(n=743) | Asthma + COVID-19<br>Hospitalized<br>(n=263) | OR (95% CI)<br>*P* value * |
| --- | --- | --- | --- | --- |
| *Age* |  |  |  |  |
| Years<br>(mean ± SD) | 55 ± 20 | 52 ± 20 | 63 ± 18 | <.001 |
| *Sex* |  |  |  |  |
| Female (%) | 66 | 67 | 61 | 0.76 (0.57 – 1.02)<br>0.07 |
| *Tobacco use (%)* | 7 | 7 | 6 | 0.69 (0.37 – 1.29)<br>0.29 |
| *Comorbidities* |  |  |  |  |
| Arterial<br>Hypertension<br>(%) | 39 | 34 | 54 | 2.27 (1.71 – 3.03)<br><.001 |
| Dyslipidemia<br>(%) | 27 | 21 | 38 | 2.31 (1.70 – 3.13)<br><.001 |
| Diabetes<br>Mellitus (%) | 15 | 13 | 21 | 1.76 (1.22 – 2.54)<br>0.002 |
| Obesity (%) | 13 | 12 | 18 | 1.60 (1.09 – 2.35)<br>0.02 |

\* Non-hospitalized patients vs. hospitalized patients

The proportion of patients with asthma using inhaled corticosteroids (ICS) was significantly lower in individuals requiring hospital admission (48.3% vs 61.5%) respectively, with an OR (95% CI) of 0.58 (0.44 - 0.77). The most common diagnosis in hospitalized patients was pneumonia (91% of patients; n = 239), with great variability of radiological expression. Different diagnoses associated with respiratory failure but with normal lung radiology (data not shown) were found in 9% (n = 24) of patients.

Regardless of previous ICS or bronchodilator use, a total of 865 patients in the study population were being treated with biologics during the study period, due to previous poor control of the disease (**Table 3**). Biologics-treated patients showed a high frequency of rhinitis and polyposis (50% in both cases), and a greater number of bronchospasm episodes before treatment onset with biologics. Despite increased severity and comorbidity of symptoms at the ear, nose, and throat (ENT) level, the need for COVID-19-related hospital admission in biologics-treated patients with asthma was relatively marginal (0.23%; 0.03 – 0.83). This proportion markedly differs from the observed in both the general population and in the population of patients with asthma not being treated with biologics; in both cases 26% of patients required hospitalization due to COVID-19. Only one patient undergoing treatment with biologics died; he was a 52-year-old male with high blood pressure, diabetes mellitus, and dyslipidemia.

**Table 3.**
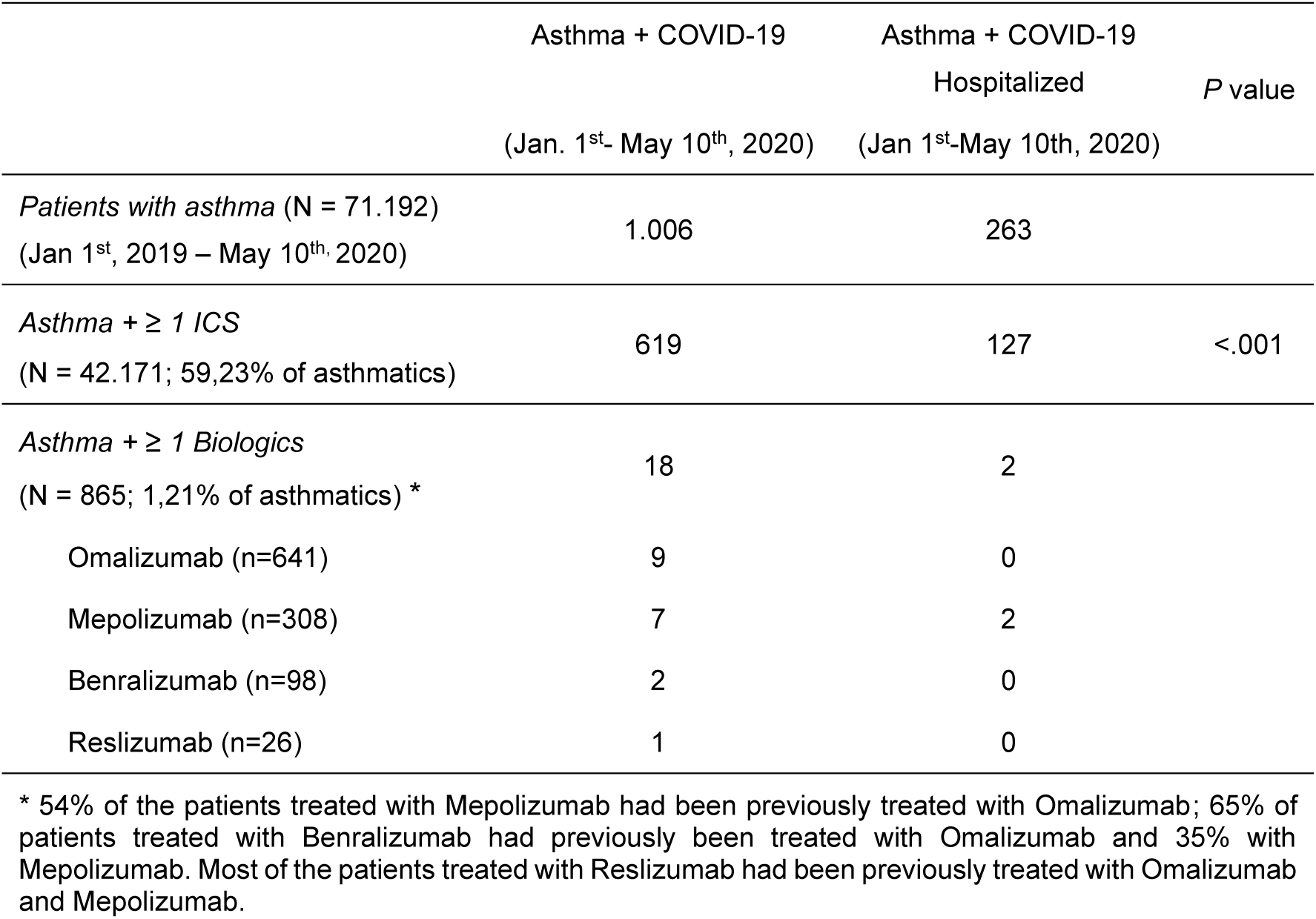
Treatment of patients with asthma and COVID-19: ICS and biologics in hospitalized vs. non-hospitalized patients.

|  | Asthma + COVID-19<br>(Jan. 1<sup>st</sup>- May 10<sup>th</sup>, 2020) | Asthma + COVID-19<br>Hospitalized<br>(Jan 1<sup>st</sup>-May 10th, 2020) | P value |
| --- | --- | --- | --- |
| *Patients with asthma* (N = 71.192)<br>(Jan 1<sup>st</sup>, 2019 – May 10<sup>th</sup>, 2020) | 1.006 | 263 |  |
| *Asthma + ≥ 1 ICS*<br>(N = 42.171; 59,23% of asthmatics) | 619 | 127 | <.001 |
| *Asthma + ≥ 1 Biologics*<br>(N = 865; 1,21% of asthmatics) * | 18 | 2 |  |
| Omalizumab (n=641) | 9 | 0 |  |
| Mepolizumab (n=308) | 7 | 2 |  |
| Benralizumab (n=98) | 2 | 0 |  |
| Reslizumab (n=26) | 1 | 0 |  |

\* 54% of the patients treated with Mepolizumab had been previously treated with Omalizumab; 65% of patients treated with Benralizumab had previously been treated with Omalizumab and 35% with Mepolizumab. Most of the patients treated with Reslizumab had been previously treated with Omalizumab and Mepolizumab.

Compared with information from patients with asthma available since January 2019, the data collected during the study period (January 1^st^ to May 10^th^, 2020) show that COVID-19 significantly increased in-hospital mortality in this population (0.54% vs 2.29%), with an associated OR (95% CI) of 4.35 (2.84 - 6.66). For both periods, in-hospital mortality mainly affected elderly patients, with an average age (±SD) of 76 (±12) years in patients with both asthma and COVID-19, and 78 (±17) in COVID-19-free patients with asthma, *P* <.001); most of these patients were women in both study periods (61% and 71%, respectively, and had previously diagnosed comorbidities (**Table 4**). On the other hand, the age distribution (mean ± SD) of patients without asthma who died from COVID-19 was 79 ± 11 years, and 63% (n = 296) of them were male.

**Table 4.** In-hospital mortality in asthmatic patients with / without COVID-19

|  | Asthma with<br>COVID-19 | Asthma without<br>COVID-19 | OR (95% CI)<br>*P* value |
| --- | --- | --- | --- |
| *Age* |  |  |  |
| Years<br>(mean ± SD) | 78 ± 17 | 76 ± 17 | <.001 |
| *Sex* |  |  |  |
| Female (%) | 61 | 71 | 0.64 (0.56 – 0.73) |
| *Mortality (%)* | 2.29 | 0.54 | 2.29 (4.35 – 6.66) |

## DISCUSSION

The WHO declared the COVID-19 outbreak a global pandemic on March 11, 2020. Ever since, clinicians worldwide have been particularly concerned about the impact of patients’ preexisting chronic conditions (particularly lung and cardiovascular diseases) on the course of this new disease. Whereas high blood pressure and diabetes have been closely associated with increased frequency of cases and severity of COVID-19, care-related data suggest that COVID-19 has not affected patients with asthma to nearly the same extent (13,21-23).

To shed light on this question, we have applied big data analytics and AI to analyze the clinical information of a large cohort of patients with asthma and COVID-19. Big data applications in healthcare (specifically the use of new technologies to manage and extract the complex data generated in large volumes of EHRs) is an inescapable reality (24). Crucially, most of the information contained in EHRs is found in an unstructured form (e.g., as free-text narratives or clinical notes). The use of big data analytics and cutting-edge methods in the realm of AI (i.e., NLP, machine learning) now allows for the extraction and analysis of this valuable information in real time. The software use in the present study enable the rapid evaluation of the main indicators of various clinical processes while avoiding the selection biases that occur in audit or case series studies, where only specialized centers and medical professionals participate in the study. Importantly, the Spanish Autonomous Community of Castilla-La Mancha has different features that optimize the use of this technology to analyze the clinical impact of COVID-19 on patients with asthma. First, this region has been one of the most affected by the pandemic in Spain, which in turn has been one of the most hard-hit countries in Europe. Second, it has a good EHR system, which has been standardized and is shared across all five provinces. Finally, the SAVANA *Manager*^*®*^ tool is widely available in the region, allowing access to large amounts of clinical information (25).

In the present study, we analyzed clinical data from the largest population of asthma patients published to date (n = 71,192); 1,006 of these patients were diagnosed with COVID-19. The study period was January 1, 2019-May 10, 2020. Although the system allows us to analyze data from 2011 onwards, we selected this temporal cut-off to include asthmatic patients with updated follow-up information and with the active form of the disease.

The proportion of patients with both asthma and COVID-19 during the study period was 1.41%, which is markedly higher than the 0.86% observed in the general population. Although these data show a higher frequency of COVID-19 in patients with asthma, the manifestation of the disease in this clinical population was not particularly severe, with low rate of hospital admissions. In addition, this proportion is lower than the reported for patients with other chronic diseases. Some of the reasons that may explain this phenomenon include remission of seasonal influenza, lack of exposure to environmental factors, greater monitoring of hygiene measures during lockdown in these patients, the significant reduction in air pollution during this period, and/or better control of the disease by improving adherence to treatment due to fear of worsening symptomatology. This trend was already observed since the initial phases of the pandemic in patient populations with other respiratory diseases such as COPD (26).

Comorbidities play a major role in the manifestation of COVID-19-related complications. In our study, the manifestation of COVID-19 in patients with asthma was favored by older age, male sex, and the presence of several comorbidities. High blood pressure, dyslipidemia, diabetes, and obesity were the main risk factors for hospital admission due to poor prognosis. The lower risk associated with rhinitis and eczema is consistent with previous observations that allergic sensitization in asthma is linked to lower expression of ACE2 receptors in both upper and lower respiratory airways, suggesting a potential protective effect (27). As previously observed in the general population, mortality due to COVID-19 in patients with asthma mainly occurred in the elderly.

The possibility that different therapeutic options in patients with chronic respiratory diseases affects the incidence and prognosis of COVID-19 has been a matter of intense debate. As for asthma, it has been suggested that the use of ICS might yield a protective effect against COVID-19 (28,29). Although an antiviral effect has been described in rhinovirus-induced exacerbations, these results are highly controversial. Peters et al (30) showed an association of ICS use and reduced expression of both ACE2 and TMPRSS2 receptors, thus implying that ICS may reduce the risk for SARS-CoV-2 infection and decrease COVID-19-related morbidity. Although our data cannot answer this question in a conclusive manner, they show that the proportion of patients who used ICS was significantly reduced in asthmatics who required hospitalization due to COVID-19. These findings are consistent with other studies showing that a combination of glycopyronium, formoterol, and budesonide prevents the replication of HCoV-229E (via inhibition of receptor expression and/or endosomal function), and that these drugs modulate infection-related inflammation in the respiratory tract (31). In our study, the three cortico-dependent patients in our study diagnosed with COVID-19 did not die from it; however, there is no experimental evidence to date regarding the effect of systemic corticosteroids in patients with asthma and COVID-19.

Whether treatment with biologics in patients with asthma impacts SARS-CoV-2 infection or the incidence and prognosis of COVID-19 also remains unknown. In this context, there is no evidence supporting that either omalizumab or other drugs that suppress eosinophils directly modulate viral processes in patients with asthma (32,33). In our study, we identified a total of 865 patients treated with biologics. Among these, two patients required admission and only one died from COVID-19; this patient, however, had other comorbidities in addition to severe asthma, which may have contributed to his poor clinical outcome. Overall, our results support the safety of these drugs for the treatment of asthma in patients diagnosed with COVID-19. As with ICS use, our data suggest that biologics might be associated with a protective effect on the clinical course of these patients. However, we cannot rule out that the aforementioned favorable factors also contributed to a better disease prognosis.

Our findings are consistent with a recently reported clinical series of 220 asthmatic patients with COVID-19 in Chicago and surrounding Illinois suburbs, where asthma was not associated with an increased risk of hospitalization after adjusting for age, sex, gender, and comorbidities (RR of 0.96 [95%CI: 0.77-1.19]) (34). However, ICS use was associated with a non-statistically significant increased risk of hospitalization (RR of 1.39 [95%CI: 0.90-2.15]); in the same report, however, the interpretation of the effects of combined use of ICS and long-acting beta agonists (LABA) and levels of care in these patients was not straightforward. These results warrant further research in patients with asthma and other clinical populations.

## Strengths and Limitations

The strengths of the present study include immediacy, large sample size, and real-world evidence. In addition, our results must be interpreted in light of the following limitations. Frist, the main limitation of this type of studies is perhaps the lack of documented information. In Castilla-La Mancha, the digitalization of clinical records has been optimal since 2011. Not only the EHRs system is homogenous throughout the region, but its use has been universal for the past five years. Second, unlike classical research methods, reproducibility is not generally considered in big data studies, since the latter involve large amounts of information collected from the whole target population. Because we exclusively analyzed the data captured in EHRs, the quality of the results reported for some variables is directly tied to the quality of the clinical records; in many cases, EHRs may be partially incomplete and not capture all the relevant clinical information from a given patient. Third, because this study was not designed to collect variables in a strict, *a priori* fashion, there were some variables that were not properly documented and were therefore not analyzed. Finally, our study sample comprised COVID-19 cases confirmed by both PCR/serological tests and clinical criteria (i.e. symptomatology, imaging, and laboratory results). Of note, PCR and other rapid laboratory tests for the detection of SARS-CoV-2 were not routinely used in Spain during the onset of the pandemic. The decision to include clinically diagnosed COVID-19 cases is further supported by recent data questioning the clinical validity and sensitivity of symptom- and image-based identification of patients with COVID-19, especially in the early stages of the disease (35-37).

## Conclusion

We conclude that *a)* the frequency of SARS-CoV-2 infection has been low in patients with asthma, although higher than in the general population, *b)* the increased risk for hospitalization due to COVID-19 in patients with asthma is largely associated with age and related comorbidities; mortality mainly affected elderly patients, *c)* ICS showed a safe profile; compared to asthmatic patients who required hospitalization due to COVID-19, a significantly higher percentage of non-hospitalized patients used ICS, and *d)* although biologics-treated patients with asthma typically present with the most severe manifestations of the disease, the number of COVID-19-related admissions and mortality in these patients was strikingly low, thus suggesting a protective effect associated with the use of these therapeutic agents.

## Data Availability

Data not publicly available due to legal restrictions

## ONLINE APPENDIX

**eTable 1. Search terms used to identify patients with bronchial asthma**

- Unstable asthma
- Adult-onset asthma
- Intrinsic asthma
- Asthma attack / Asthma exacerbation
- Acute asthma
- Exercise-induced asthma
- Childhood asthma
- Asthmatic bronchitis
- Cough-variant asthma
- Allergic asthma
- Mild asthma
- Moderate asthma
- Occasional asthma
- Severe asthma
- Chemical-induced asthma
- Substance-induced asthma
- Intermittent asthma
- Seasonal Asthma
- Occupational Asthma
- Chronic obstructive airway disease with asthma
- Asthma with irreversible airway obstruction
- Untreated Asthma
- Persistent asthma
- Induced asthma
- Neutrophilic asthma
- Cortico-dependent asthma

